# Implementation of Tuberculosis Monitoring Encouragement Adherence Drive (TMEAD) for improving treatment adherence amongst drug-resistant Tuberculosis patients in Ahmedabad City, Gujarat

**DOI:** 10.1101/2023.10.18.23297189

**Authors:** Poonam Travedi, Devang Raval, Danish Malik, Somen Saha, Deepak Saxena, Nishad Halkarni, Rahul Doshi, Sukan Rajpurohit, Raghavendra Rao, Madhav Joshi

**Author notes:** **Address for Correspondence**, Dr. Somen Saha, Professor, Indian Institute of Public Health Gandhinagar Gujarat, Email id.

## Abstract

**Background:** Treatment Adherence among Tuberculosis (TB) patients is a critical challenge globally and in India. Digital technology has appeared as a tool for providing patient centric monitoring support to improve TB patients’ drug adherence. However, there is a paucity of evidence on the acceptability and effectiveness of such devices for enhancing adherence among drug-resistant TB (DRTB). The present study aims to document the feasibility of using the Tuberculosis Monitoring Encouragement Adherence Drive (TMEAD) and treatment adherence amongst DRTB patients.

**Methods:** A longitudinal follow-up study of DRTB patients was conducted in Ahmedabad. A total of 22 Tuberculosis Units (TUs) from Ahmedabad’s rural and urban regions were included in the study based on the high load of DR-TB patients. Two hundred patients were enrolled per the inclusion criteria, and the TMEAD device was deployed to the enrolled patients and followed up monthly for six months to document drug adherence and various challenges.

**Result:** More than 80.5% of the patients used the device, and the point drug adherence was 92% among the patients who used the device. About 19.5% did not use the device, and the reasons for the non-use of the device include non-functional devices and alarms, charging, and incomplete refilling with prescribed drugs. The other significant issues include the social stigma regarding the disease and the fear of disclosure of TB.

**Conclusion:** The present study revealed that the acceptability of TMEAD and patient reported drug adherence were high. However, there were various disease and device related challenges. The implementation of TMEAD can be improved through the design modification. Further large-scale research is required to document the effectiveness of the device and scale up.

## Introduction

Tuberculosis (TB) is one of the most common causes of death worldwide. In 2021, approximately 10.6 million individuals worldwide contracted tuberculosis (TB), reflecting a 4.5% increase from the 10.1 million cases reported in 2020(1). According to the India TB report, India had an estimated 2.69 million tuberculosis (TB) cases, representing 25% of global TB cases. (2). Further, Drugresistant TB in India is also a major public health problem in India; as per the global TB report, India accounted for 26% of the total global MDR cases in 2021(1,3).

To achieve the Sustainable Development targets of TB by 2025, the government of India took various initiatives to combat TB. The National Strategic Plan (NSP) for the period 2017-2025 envisions an India free of tuberculosis (TB), aiming to achieve zero deaths, zero disease, and zero poverty caused by TB and incorporating new guidelines for the rapid expansion of molecular diagnostic services to address drug-resistant TB (DRTB) (4). Despite all efforts, the DRTB cases continue to rise in India. In 2022, there was a 32% increase in MDR cases detected under the National Tuberculosis Elimination Program (NTEP) compared to 2021. (2).

Treatment compliance is essential for successfully controlling and eradicating TB in India. Poor TB medication adherence also increases rates of poor outcomes, disease relapse and transmission of drug-resistant strains (5). Treatment adherence amongst Drug-resistant patients is a significant challenge for controlling and preventing the spread of drug-resistant cases. Hence, it is critical to ensure the treatment adherence of Drug-Resistant Patients. There are multiple factors affecting drug adherence, which include side effects of medicine, duration of treatment, psychosocial, stigma and health system-related challenges(6,7).

Digital adherence solutions have emerged to address traditional directly observed therapy (DOT) barriers to providing more patient-centric monitoring support(8). These technologies include cell phones, digital pillboxes, SMS reminder systems and sensors. Digital adherence technologies (DAT) can potentially improve and remotely monitor medication adherence(9). Since 2015, the government has implemented innovative technology like 99 DOTS, a cell phone-based Digital Adherence Technology (DAT), to improve TB patients’ monitoring and treatment compliance(10). However, there is limited evidence on using such technology to enhance treatment compliance amongst TB patients, especially drug-resistant TB cases.

### Tuberculosis Monitoring Encouragement Adherence Drive (TMEAD)

Tuberculosis Monitoring Encouragement Adherence Drive (TMEAD), an IOT-enabled device, was developed by Sense Dose technology. TMEAD is the technology-based solution to enhance adherence to the NTEP regimen by daily regimen monitoring, avoiding interruption in the treatment, reducing defaulters, analyzing patients’ data and reducing the burden of TB Health Visitors (TBHV) in maintaining records and tracking patient adherence. A physical device equipped with an integrated software-linked mobile network reminds dispenses, and senses a patient’s adherence to the treatment regime. A web-based application providing real-time monitoring with daily updates and patient analytics to peripheral health institutions (PHI). A mobile application provides instant updates and a quick view for the TB Health Visitor (TBHV) when they are on the field.

A quasi-experimental study was conducted in Maharashtra state to explore adherence and cost-effectiveness of TMEAD compared to the standard care among patients with drug-sensitive Tuberculosis (DSTB). There was high treatment adherence in the intervention group (99%) than in the control group (90%) and the device was well-accepted by the drug-sensitive patients(11). However, the acceptability and feasibility of such a device for drug-resistant TB with more complex and prolonged treatment remain unexplored. Hence, based on a further recommendation from the Central TB Division, a longitudinal study was conducted in Ahmedabad, Gujarat, to explore the feasibility of using this device amongst Drug Resistance TB (DRTB). The objective of the present paper is to document the feasibility of using the TMEAD device amongst DRTB patients and to document the treatment adherence of DRTB patients who used the TMEAD device. The various challenges of using TMEAD devices amongst DRTB patients were also recorded.

## Methods

### Study Design

A longitudinal follow-up study was conducted to ascertain treatment adherence amongst the patients who used the TMEAD device.

### Study Settings

The present study was conducted in the Ahmedabad district of Gujarat from Jan-Oct 2022. Both urban (AMC) and rural areas (Ahmedabad district) were included in the study to have better representation. 17 out of 17 TUs of Ahmedabad Municipal Corporation and five from Ahmedabad district with high DRTB patient loads were chosen for the study. The study included newly diagnosed DRTB patients (per the NTEP definition), those aged ≥16 years and willing to participate. As the study’s objective is to document the adherence amongst the patients, only the patients on all oral, more extended treatment regimens and a Shorter MDR treatment regimen (9 months), with at least six months remaining on their DRTB treatment regimen, were included in the study. At the same time, seriously ill patients in the extended IP phase who were not willing to participate in the study were excluded. 200 DRTB patients from selected TUs were purposively selected and enrolled based on the inclusion criteria.

### Recruitment of the participants and data collection

Before the rollout of the TMEAD, The TB-HVs and researchers were trained on the appropriate use of the TMEAD device. TB -HVs dispensed medication refills in the TMEAD device for enrolled patients. First, the enrollment of the patients was done based on the inclusion criteria, and then the patients were followed up monthly for six months. A close-ended questionnaire was developed based on the literature review administered to the patients during enrollment. Before deployment of the devices, the researchers explained the advantages and how to use the device to the enrolled patients. The patient’s written informed consent was taken before the deployment of the device and interview. Researchers from the IIPHG team interviewed the patients at the selected TUs. The sociodemographic characteristics of the patients, family and previous treatment history were documented. Although 200 patients were enrolled, the devices were not deployed in six patients as five refused, and one died before the deployment.

After the deployment of devices, the medicine was refilled in the TMEAD box by TB-HVs of respective TUs at the follow-up time, and they were paid an additional incentive for device refilling. During their follow-up visit, the researchers followed up with 194 eligible patients based on the deployment date till six months after TMEAD device deployment. Household visits were also done if the patient failed to visit the centre. During the follow-up visit, information on treatment adherence was collected, and if the patients could not be contacted for more than two visits, they were labelled as lost to follow-up. In the present study, the operational definition for adherence was patients on TB treatment and completing 80% of the prescribed doses for treatment.

In addition to compliance, patient experiences like device-related technical problems, device usefulness, the availability of DRTB medicines, and the status of monthly medicine refilling were also documented during each follow-up phase. Based on the treatment start date, the patient’s status at the time of each follow-up was documented. The selected TUs also received the treatment adherence details from the TMEAD device. However, it was not used for the present study. Treatment outcomes were classified as treatment completed, on treatment, defaulters and lost to follow-up (if any) per the NTEP.

### Data analysis

Data was entered and cleaned in Excel, and a descriptive analysis was done using SPSS version 21.

## Result

### Sociodemographic Profile of the study participants

Table 1 shows the sociodemographic characteristics of the patients enrolled in the study. Of 200 patients enrolled in Ahmedabad-Gujarat, 49% were male, and 51% were female. The average age of patients was 32.8 ± 31.5 SD years, and most were Hindu (75.5%). Approximately 89.5% of study participants were literate. Most patients (69.5%) were married, followed by unmarried (38%) and divorcees (1.5%). Sixty-three per cent of the patients were unemployed, and the family size ranged from 1 to 25 family members. The average annual household income of the study participants was Rs. 2,03,870 (Rs. 25000 - 7,00,000) in Ahmedabad.

**Table 1:** Sociodemographic characteristics of the study participants (n= 200).

| **Variables** | **Gujarat N (%)** |
| --- | --- |
| **Gender** |  |
| Male | 98(49.0) |
| Female | 102(51.0) |
| **Age group (in years)** |  |
| 16 to 20 years | 48 (24) |
| 21 to 40 years | 96(48) |
| 41 to 60 years | 49(24.5) |
| 61 years and more | 7(3.5) |
| **Age in Mean Years** | $32.8 \pm 31.5$ |
| **Caste** |  |
| General/Open | 94(47) |
| S.C/S. T | 33(16.5) |
| OBC | 73(36.5) |
| **Religion** |  |
| Hindu | 151(75.5) |
| Muslim | 47(23.5) |
| Others | 2(1) |
| **Education** |  |
| Literate | 179(89.5) |
| Illiterate | 21(10.5) |
| **Marital status** |  |
| Unmarried | 76(38) |
| Married | 121(60.5) |
| Divorced/Widowed/Separated | 3(1.5) |
| **Present Occupation** |  |
| Unemployed | 126(63) |
| Employed | 74(37) |
| Average Family Size of HH | 3 (Min 1 to Max. 25) |
| Average HH Income (yearly) | Rs. 2,03,870 (min-Rs.25000-max-7,00,00) |

**Abbreviation:** HH: House Hold, SC: Schedule cast, ST: Schedule Tribe

Of a total of 200 patients enrolled, 84.5% were on Longer oral regimens in Ahmedabad, Gujarat, while 13.5% and 2% were on shorter MDQ/Bdq and all oral H monopoly, respectively. About 21.5% reported comorbidities like Diabetes and Hypertension. Of the total 200 patients, the majority (90.5%) had pulmonary while 9.5% had extrapulmonary TB. Most patients (57%) were diagnosed in 2021, while the remaining 41% and 2% were diagnosed in 2022 and 2020, respectively.

### Follow-up details of the study participants

The patients were contacted after one month of device deployment; hence, the deployment date varies from patient to patient and between different months. As presented in Table 2, out of 194 patients eligible for the 1^st^ follow-up, 170 could be contacted; the remaining 24 could not be contacted due to multiple reasons like migration, patient not reachable, death, the patient not wanting to talk and stigma. For the second follow-up, out of 194 patients, 14 were not eligible for follow-up as three patients died, and 11 returned the device. Hence, the remaining 180 were eligible for follow-up. Similarly, the eligible patients were followed up during the rest of the follow-up, while those who returned the device and died were excluded. During the last follow-up, 137 were eligible for follow-up, out of which 136 were contacted.

**Table 2.** Monthly follow-up status of the enrolled patients.

| **Follow-up status** | **FU1** | **FU2** | **FU3** | **FU4** | **FU5** | **FU6** |
| --- | --- | --- | --- | --- | --- | --- |
| Eligible for follow-up | 194 | 180 | 164 | 160 | 150 | 137 |
| Follow-up done | 170 | 163 | 157 | 159 | 149 | 136 |

**Table 3:** Use of TMEAD Device during Follow-up.

| **Follow-ups** | **FU1** | **FU2** | **FU3** | **FU4** | **FU5** | **FU6** | **Avg. %** |
| --- | --- | --- | --- | --- | --- | --- | --- |
| **No. of patients Eligible** | **n=170** | **n=163** | **n=157** | **n=159** | **n= 149** | **n= 136** |  |
| Using device | 138 (81.2) | 130 (79.8) | 124 (79.0) | 132 (83) | 118 (79.2) | 110 (80.9) | 80.5% |
| Not Using | 21 (12.4) | 20 (12.3) | 29 (18.5) | 18 (11.3) | 23 (15.4) | 23 (16.9) | 14.5% |
| Returned the device | 11 (6.5) | 13 (8.0) | 4 (2.5) | 9 (5.7) | 8 (5.4) | 3 (2.2) | 5.04 % |

**Table 4:** Details of treatment compliance and reasons for the missed dose.

|  | **FU1** | **FU2** | **FU3** | **FU4** | **FU5** | **FU6** | **Overall** |
| --- | --- | --- | --- | --- | --- | --- | --- |
| **Variables** | (n=159) | (n=145) | (n=131) | (n=133) | (n=120) | (n=109) |  |
| **Missed dose** |  |  |  |  |  |  |  |
| Yes | 7 (4.4) | 6 (4.1) | 13(9.9) | 8(6.0) | 14 (11.7) | 10 (9.2) | 7.5% |
| No | 152 (95.6) | 139 (95.9) | 118 (90.1) | 125 (94) | 106 (88.3) | 99 (90.8) | 92% |
| **Reasons for missed doses** | **n=7** | **n=6** | **n=13** | **n=8** | **n=14** | **n=10** | **Overall** |
| Personal Negligence | 3(42.9) | 1(16.7) | 5(38.5) | 3(37.5) | 8(57.1) | 4(40.0) | 24(41.4) |
| Medicine Side effects | 2(28.5) | 1(16.7) | 1(7.7) | 1(12.5) | 1(7.1) | 2(20.0) | 8(13.8) |
| Social Reasons | 1(14.3) | 1(16.7) | 2(15.4) | 1(12.5) | 0(0) | 0(0) | 5(8.6) |
| Travelling | 1(14.3) | 3(50.0) | 3(23.0) | 0(0) | 1(7.1) | 1(10.0) | 9(15.5) |
| Other* | 0(0) | 0(0) | 2(15.4) | 3(37.5) | 4(28.6) | 3(30.0) | 12(20.7) |

\*Hospitalization due to other comorbidities, strike

During the first follow-up, a total of 194 patients followed up; however, only 137 patients could be contacted during the last follow-up, while the rest of the 57 patients could not be approached, with a loss to follow-up of 29.3%. The reason for the loss to follow up includes the number not in service and migration.

### Usage of TMEAD device during various stages of follow-up

During all follow-ups, an average of 80.5 % of patients used the device regularly, while 14.5% did not use their device for a variety of reasons, including the machine not working, Alarm/Charging issue/Damaged, Refilling not completed, Died, and causes like travelling for more days and social stigma (Table-3). An average of 5.04 % of the patients returned the devices as they were uncomfortable with the device due to alarm issues like rings at any time, does n0t ring, the device not working correctly, and problems related to the device not being solved.

### Treatment Compliance of study participants

As presented in Table. 4, although 170 patients were followed up, the treatment compliance of only those patients who did not return the device (n=159) was documented. The treatment adherence was good in most patients, with point adherence of 92%. During the first follow-up, out of 159 patients, only 07 (4.4%) missed the dose. From the second follow-up onwards, 4.1%, 9.9 %, 6%, 11.7%, and 9.2% of patients missed the doses, respectively. The main reason for the missed dose was related to the personal negligence of the patients and travelling to other states. It is significant to note that device-related issues were not listed among patients who missed their treatment regimen.

The present study also documented the side effects of the medicine during each follow-up. During each follow-up, 17.1%, 14.1%, 9.6%, 10.7%, 8.1%, and 10.3% reported the side effects of medicine during the 1^st^ and 6th follow-up, respectively. The most common side effects were nausea and vomiting 24% and joint pain 12.5%.

During the last follow-up, the status of the eligible patient was also documented, which shows that about 90% of the patients were on the treatment, while 2.1% completed the treatment. Very few patients were defaulters (5.2%), while 1% were lost to follow-up.

## Discussion

The present study documented the feasibility of using the TMEAD device amongst the DRTB patients in the Ahmedabad district of Gujarat. The acceptability of the TMEAD was good amongst the enrolled participants, with more than 80% of them using the device. However, the existing studies reported the variability in acceptance of such digital pill boxes. A pilot evaluation of an electronic pill box for MDR TB patients in South Africa found a higher acceptance of the device(12). On the contrary, a study conducted in Vietnam aimed to implement a Medication Event Reminder Monitor (MERM) box among drug-sensitive patients revealed that only half of the patients used the device(13). A qualitative study conducted in two states of India to explore the acceptability of the MERM Box amongst Drug-resistant patients reported the variability in acceptability(6).

The non-usage of devices amongst the enrolled participants was 14%. The reason for non-usage includes both technical issues and personal reasons. The technical issues include improper machine functioning, damaged or uncharged adequately, and incomplete refilling. Other causes include patient deaths, travelling for more days and social stigma. Similarly, the disease-related stigma was also reported by recent studies as a barrier to usability among patients with MDR-TB (6). In the present study, an average of 5.04 % of the patients returned the devices due to device-related issues like an alarm at any time, not ringing, or inadequate device functioning.

The present study highlighted the high treatment adherence among the study participants. The point adherence to the treatment was 92% amongst the enrolled participants with a low defaulter rate, which shows that the device helps improve compliance. Similarly, a cluster randomized trial conducted in China with drug-susceptible TB patients found that digital pillboxes effectively reduce the percentage of patient months with high nonadherence(14).

Faster adoption of TMEAD technology will require significant design thinking, including:

- Medication Interactions Checks: Integrate a database of medications with their interactions in the mobile app.
- Dosage Adjustments and Reminders: Enable the system to suggest and adjust medication dosages based on user-specific needs.
- A robust mechanism for refilling and patient reporting – Establish patient-centric support where they verify their dose

To the best of our knowledge, the present study is the first qualitative study to document the use of digital pillboxes amongst Drug-resistant TB patients in India. In the present study, both the acceptability and the effect of the device on treatment adherence amongst DRTB patients were also documented. Secondly, the study was conducted amongst a large sample size and covered both urban and rural populations. However, there are a few limitations of the present study. First, the present study was longitudinal, with non-random sampling; hence, the findings cannot be generalized. The treatment compliance may be overestimated as it was captured from the patients and not by the system and verified by other means such as biomedical options.

## Conclusion

The present study revealed acceptance and feasibility of using TEMED amongst the DRTB patients. The treatment adherence amongst the patients who used the device was also good. However, some barriers hindered the device’s usability, which can be addressed through device modification and the introduction of robust mechanisms for refilling and patient reporting. At the same time, there were also disease-related issues that affected treatment compliance that may restrict the use of the device, which requires more efforts towards generating awareness about TB amongst the community and should be considered for implementation of the device on a larger scale. Further research is needed to generate evidence on the cost-effectiveness of the device amongst DRTB patients with a larger sample size.

## Abbreviation

HH: House Hold
SC: Schedule cast
ST: Schedule Tribe

## Acknowledgement

This research was conducted through a collaborative effort between the IIPHG and the State TB Division of the Health and Family Welfare Department, the Government of Gujarat.We extend our gratitude to Dr. Karthik Shah and Dr Tejas Shah from the Department of Health-State TB Cell in Gujarat for their invaluable support during the execution of this study.

## Ethics statement

The study was approved by the Technical Appraisal Committee for Health Technology Assessment of the Department of Health Research, New Delhi and the Institutional Ethics Committee of the Indian Institute of Public Health, Gandhinagar, vide TRC-IEC No: 02/2020-21 dated 29 May 2020. The patients/participants provided their written informed consent to participate in this study.

## Author contribution

All authors contributed equally to the development of this study. PT, DS & SS participated in the conception and design of the study protocol. DM, DR and PT involved in project administration. DM & PB analyzed the data. PT & SY finalized the analysis. PT drafted the first draft of the paper. All the authors SS and and DS critically reviewed the paper. All authors read and approved the final manuscript.

## Data availability statement

Data from this study will be available at the Indian Institute of Public Health Gandhinagar (IIPHG), India, after the completion of this study. Researchers who meet the criteria for access to confidential data are encouraged to approach Dr. Somen Saha.

